# Effect of viral replication and liver fibrosis on all-cause mortality in HIV/HBV coinfected patients: a retrospective analysis of a 15-year longitudinal cohort

**DOI:** 10.1101/2021.04.13.21255432

**Authors:** Lorenza N. C. Dezanet, Raisha Kassime, Patrick Miailhes, Caroline Lascoux-Combe, Julie Chas, Sarah Maylin, Audrey Gabassi, Hayette Rougier, Constance Delaugerre, Karine Lacombe, Anders Boyd

## Abstract

**Background:** In patients co-infected with HIV and hepatitis B virus (HBV), widespread tenofovir (TDF)-containing antiretroviral therapy (ART) has led to substantial decreases in HBV-DNA and HIV-RNA detection. However, the link between viral replication, liver fibrosis, and mortality remains unclear.

**Methods:** 300 HIV-HBV co-infected patients undergoing ART were prospectively followed. Virological and clinical data were obtained at baseline and every 6-12 months. We quantified the association between HBV-DNA, HIV-RNA, and liver fibrosis with risk of all-cause mortality using a joint longitudinal-survival model. Viral detection, viral loads, and time-averaged cumulative viral loads of HIV and HBV were modeled as three separate exposures.

**Results:** During a median 10.5 years (IQR=4.0-14.6), the proportion undergoing TDF-containing ART (baseline=18.7%, end of follow-up=79.1%) and with undetectable HBV-DNA (baseline=36.7%, end of follow-up=94.8%) substantially increased. HIV-RNA was mostly undetectable during follow-up (76.6%). 42 participants died (incidence rate=1.30/100person-years, 95%CI=0.96-1.76). The leading causes of death were non-AIDS/non-liver-related malignancies (28.6%), closely followed by liver-related (16.7%), AIDS-related (16.7%), and other (16.7%). All-cause mortality was associated with HBV-DNA viral load (adjusted-HR per log_10_IU/mL=1.41, 95%CI=1.04-1.93, *p*=0.03) or time-averaged cumulative HBV-DNA (adjusted-HR per log_10_IU-years=1.37, 95%CI=1.03-1.83, *p*=0.03), but not undetectable HBV-DNA (adjusted-HR=0.30, 95%CI=0.08-1.09, *p*=0.08). Liver fibrosis at baseline also significantly increased mortality rates (adjusted-HR=2.35, 95%CI=1.16-4.76, p=0.02). No significant association between HIV-RNA replication and mortality was observed.

**Conclusions:** Concurrent and historical HBV replication and liver fibrosis are important drivers of all-cause mortality in largely TDF-treated HIV-HBV co-infected patients, despite one-fifth of deaths being liver-related. HBV-DNA and liver fibrosis remain important prognostic indicators for this patient population.

**Key-points:** HBV-DNA levels and cumulative exposure over time increases risk of all-cause mortality in HIV-HBV co-infected patients. Fibrosis was a major determinant of mortality; however, the leading causes of death were malignancies not related to AIDS or HBV-infection.

## Introduction

Roughly 8% of individuals living with human immunodeficiency virus (HIV) are chronically co-infected with hepatitis B virus (HBV) [1]. Without effective treatment, HIV-HBV co-infected individuals are at increased risk of both liver-related and all-cause mortality when compared to HIV-positive individuals without HBV infection [2].

Treating co-infected individuals primarily involves controlling both HIV and HBV replication. Higher levels of serum of HIV-RNA have been associated with increased risk of AIDS-related morbidity and mortality in HIV-positive individuals [3]. With effective antiretroviral therapy (ART), HIV replication is suppressed and the risk of HIV-related morbidity and mortality greatly decreases [4,5]. Likewise, higher levels of circulating HBV-DNA have been strongly linked to fibrosis, hepatocellular carcinoma (HCC), and liver-related and overall death in HBV mono-infected individuals [6]. Currently available nucleoside/nucleotide analogues (NA) can suppress HBV-DNA replication, which coincides with liver fibrosis regression [12] and reduced risk of HCC [8]. Conveniently, antiretroviral agents, such as tenofovir (TDF) and tenofovir alafenamide (TAF), possess potent anti-HIV and anti-HBV activity [9,10], and are thus ideal therapeutic options for HIV-HBV co-infected individuals [11]. It could be hypothesized that reductions in both HIV and HBV replication would give way to lower incidence of both HIV- and HBV-related causes of morbidity and mortality.

Nevertheless, epidemiological studies in co-infected populations are not entirely clear on the relationship between active replication of these viruses and mortality. Recent research has demonstrated clear reductions in AIDS-related, liver-related and overall mortality in the years coinciding with widespread TDF-use [12,13]. However, incidence of HCC in TDF-treated co-infected individuals above the age of 45 remains high enough to warrant increased HCC screening [14]. Furthermore, large studies from Tanzania and Côte d’Ivoire have shown that HIV-HBV co-infected individuals, particularly when their HBV-DNA levels are high, are still at increased risk of overall mortality despite TDF-containing ART [15,16]. The principal causes of death in these studies appeared to be related to HIV-related illness or invasive bacterial infections. The common limitation shared across all the studies conducted thus far is the lack of consistently collected data on both HIV- and HBV-replication.

The aim of the present study was then to describe detailed causes of mortality in a cohort of HIV-HBV co-infected patients followed for up to 15 years with highly effective anti-HIV and anti-HBV treatment. We further develop our analysis by exploring the effect of HBV-DNA and HIV-RNA replication over time on all-cause mortality in this study population.

## Patients and Methods

### Study population

We analyzed HIV-HBV co-infected patients from the French HIV-HBV Cohort Study. Briefly, this was a closed, prospective, longitudinal cohort study including 308 HIV-positive patients with chronic HBV infection from four centers located in Paris and Lyon, France. Patients were included if they had an HIV-positive serological result confirmed by western blot and HBsAg-positive serological results for >6 months. Participants were recruited in 2002-2003 and followed every 6-12 months until 2017-2018. The cohort design and procedures are described elsewhere [17]. For this analysis, we included patients who had at least two consecutive visits and available information on vital status.

### Ethics

All patients provided written informed consent to participate in the study and the protocol was approved by an appropriate Hospital Ethics Committee (Paris, France) in accordance with the Helsinki Declaration.

### Data collection

Demographic information was collected at study inclusion. Medical history on antiretroviral and anti-HBV treatments, alcohol consumption and the presence of comorbidities, including diabetes, cardiovascular disease (CVD), renal and other liver diseases, were collected at study entry and at each follow-up visit.

Laboratory data were collected at study entry and at each follow-up visit. HIV-RNA viral load (VL) and HBV-DNA VL were measured using a commercial PCR-based assays and CD4^+^ cell count using standard methods. Antibodies to hepatitis C virus (HCV) and hepatitis D virus (HDV) were measured with an ELISA-based assay and if positive, serum HCV-RNA and/or HDV-RNA was quantified by either commercial PCR-based assay (for HCV-RNA) or in-house assay (for HDV-RNA).

Liver fibrosis was assessed at study entry and each yearly interval using the FibroTest® [18]. METAVIR equivalents for HIV-HBV coinfected patients were used to grade liver fibrosis (F2=0.48-0.58, F3=0.59-0.73, F4≥0.74) [19].

### Mortality outcome assessment

Deaths observed during follow-up, along with the underlying cause of death and date of death, were reported by the treating physician. To obtain vital status for patients lost to follow-up (LTFU), a trusted third party (Inserm U1018) was requested to link data from the French HIV-HBV cohort to a national identification registry (*Répertoire national d’identification des personnes physiques*). For individuals reported as deceased, the cause of death was then obtained by a separate trusted third party (Centre d’épidémiologie sur les causes médicales de décès, CépiDc), linking data from the French HIV-HBV cohort to a national registry of death certificates and death notifications. Both registries are managed by the *Institut National de la Statistique et des Etudes Economiques*. Causes of death were classified by ICD-10 codes. We recategorized causes of death as liver-related, AIDS-related, non-AIDS and non-liver-related malignancies, CVD-related, other, or unknown.

### Statistical analysis

Baseline was defined as the date of study entry. Follow-up began at baseline and continued until the date of death or of the last study visit.

We assessed treatment efficacy with undetectable HBV-DNA VLs (<60 IU/mL) and HIV-RNA (<50 copies/mL). The extent of replication was assessed with HBV-DNA (log_10_ IU/mL) and levels HIV-RNA (log_10_ copies/mL). Finally, the historical extent of replication was assessed with time-averaged, cumulative copy-years over follow-up time (log_10_ copy-years_TAVG_), as detailed elsewhere [20].

To analyze the contribution of HIV and HBV replication on mortality during follow-up, we simultaneously modeled (i) HBV replication, (ii) HIV replication, and (iii) all-cause mortality. We carried out a generalized, multivariate, joint longitudinal-survival model approach by which the link between these outcomes could be taken into account. First, we ran two submodels (on HBV-DNA and HIV-RNA) for three separate sets of replication outcomes (detectable VL, log-transformed VL, and log_10_ copy-years_TAVG_). The probability of having a detectable VL was assumed to be Bernoulli-distributed and modeled using logistic regression, while mean log-transformed VL and mean log_10_ copy-years_TAVG_ were assumed to be continuous Poisson-distributed and modeled using Poisson regression. We adjusted, in the model regressing HBV-DNA, for HBeAg serostatus at baseline and cumulative tenofovir use (as a cubic-spline function delimited by 4 knots) and, in the model regressing HIV-RNA, for squared CD4^+^ cell count and HIV treatment era (2002-2007, ≥2008). Second, the hazards of death were assumed to have an exponential survival function and were estimated using a parametric survival model. We included the two submodels of HBV-DNA and HIV-RNA to estimate the hazards ratio (HR) and 95% confidence intervals (CI) of increasing expected probability of detectable VL, expected mean log-transformed VL, or expected mean log_10_ copy-years_TAVG_ on all-cause mortality. The survival model also included age, previous history of an AIDS-defining illness and level of fibrosis at study entry (F0-F1-F2 and F3-F4) as covariates, selected from previous analysis as described in the Supplementary Materials. We included a random-intercept across all models to account for between-patient variance at baseline, while the random-intercepts were constrained at 1 for the two submodels. Parameters from the three models were jointly estimated via maximum likelihood using the ‘merlin’ program in STATA [21].

All statistical analyses were performed using STATA (v15.1; College Station, Texas, USA). Significance was defined as a *p* value <0.05.

## Results

### Description of the study population

Of the 308 cohort participants, 8 were excluded as they did not have at least two visits (*n*=7) or had no information on vital status (*n*=1). Of the 300 included participants, most were male (84.0%), with a median age of 40 years (IQR=35-45) at study entry. Participants had a median CD4+ count of 400/mm^3^ (IQR=268-557) and 160 (53.5%) had undetectable HIV-RNA. More than half of participants (52.0%) were HBeAg-positive and HBV-DNA was detectable in 63.6% of them. Of the 281 patients (94.9%) with previous lamivudine (LAM) exposure, median LAM duration was 3.5 years (IQR=1.4-5.5) at study entry and 90 (32.0%) had baseline LAM resistance mutations.

Patients were followed for a median 10.5 years (IQR=4.0-14.6), with a maximum follow-up of 15.7 years, which totaled 2934.5 person-years of follow-up. The proportion of patients actively undergoing ART was high at study inclusion (90.0%) and rapidly evolved to 100% from the first year and until the end of the follow-up. Consequently, improvements in CD4^+^ cell counts (*p* for trend <0.001) were observed over time (Figure 1A). In addition, the proportion of patients actively undergoing TDF-based ART increased from 18.7% at baseline to 40.1% at the first year and 79.1% at the end of the follow-up (Figure 1B, *p* for trend <0.001).

**Figure 1.**
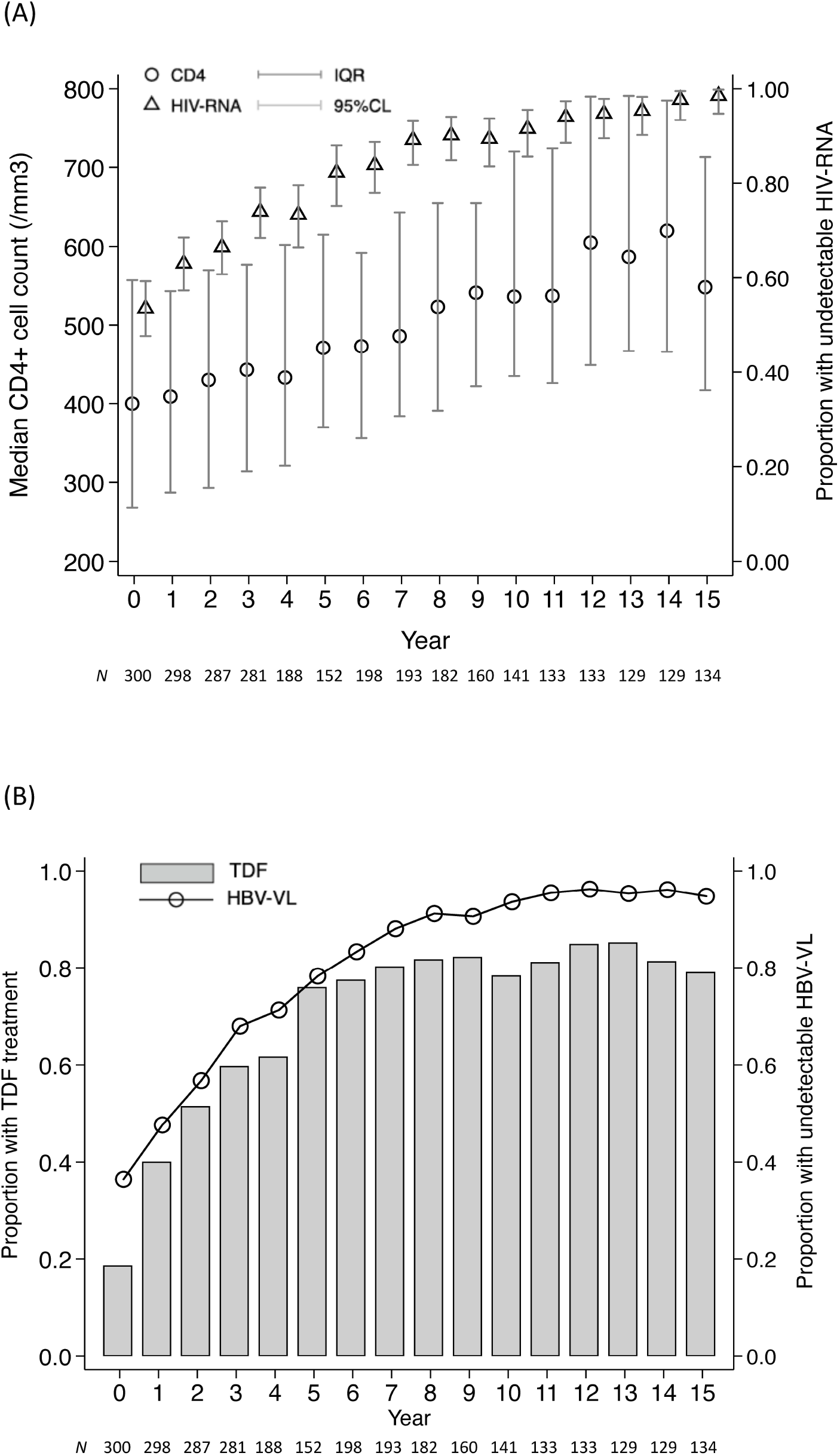
Evolution of HIV, hepatitis B virus (HBV), and antiviral treatment against hepatitis B virus over time. The number of patients continuing follow-up, as divided in yearly intervals, are provided at the bottom of each figure. In (A), undetectable HIV-RNA and median (IQR) CD4+ T-cell counts are displayed for each year. In (B), the proportion of patients with undetectable HBV-DNA viral loads, alongside the proportion undergoing antiviral therapy containing tenofovir are given for each year. Abbreviations: CI, confidence limit; HIV, human immunodeficiency virus; IQR, interquartile range.

### Description of all-cause mortality

42 deaths (cumulative incidence=14.0%; 95%CI=10.3%-18.4%) occurred after a median 6.2 years (IQR=3.4-7.9) of follow-up (incidence=1.43/100 person-years).

As shown in Table 1, the most common causes of death were non-AIDS and non-liver related malignancies (*n*=12 [28.6%]; 0.41/100 person-years), liver-related (*n*=7 [16.7%]; 0.24/100 person-years), AIDS-related (*n*=7 [16.7%]; 0.24/100 person-years) and CVD-related (*n*=6 [14.3%]; 0.20/100 person-years). HCC and hepatic failure accounted for most liver-related deaths (*n*=4 and 1, respectively). 7 patients (16.7%; 0.24/100 person-years) died from others causes of death, while for three (7.0%), the cause of death was unknown (Table 1).

**Table 1.**
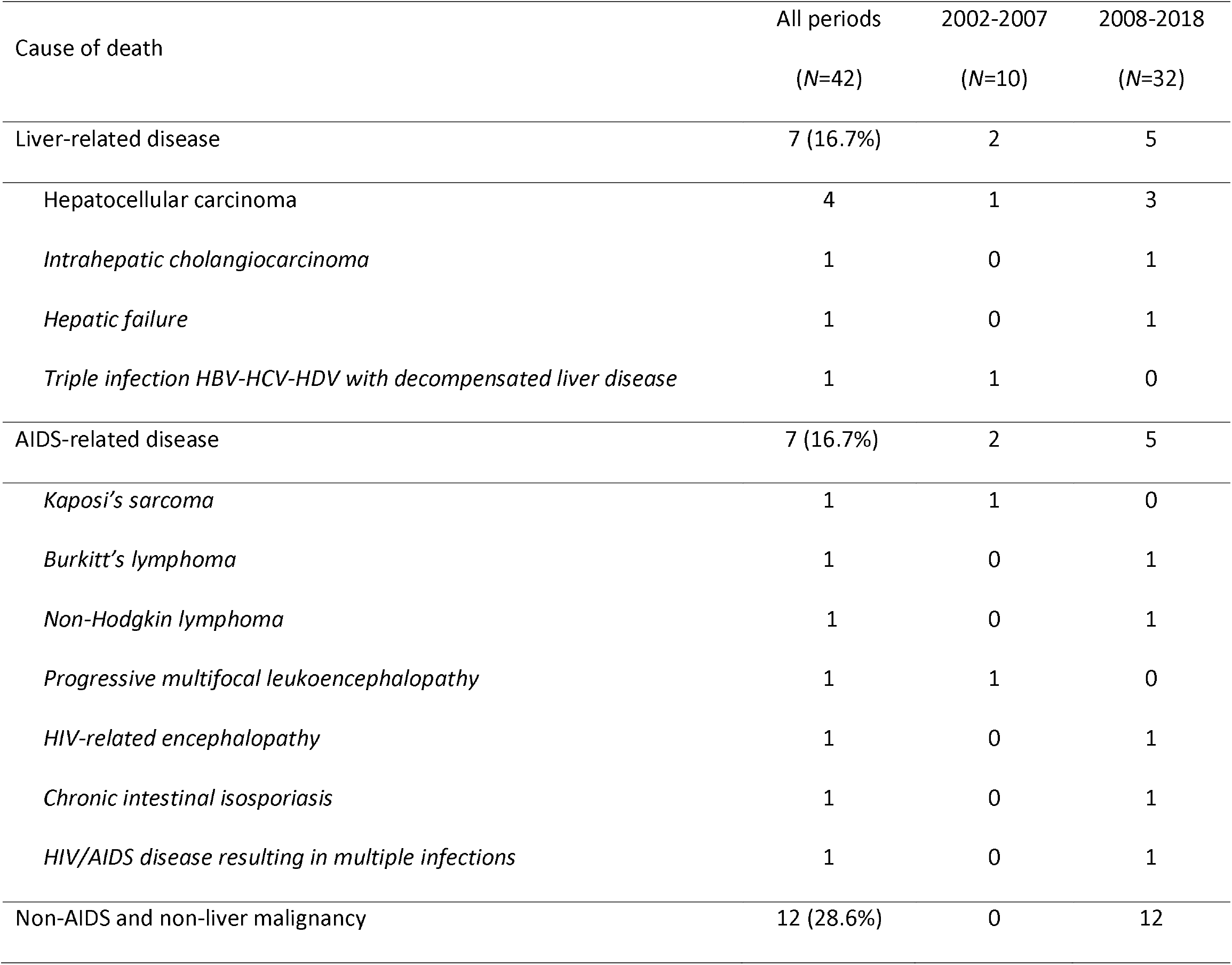
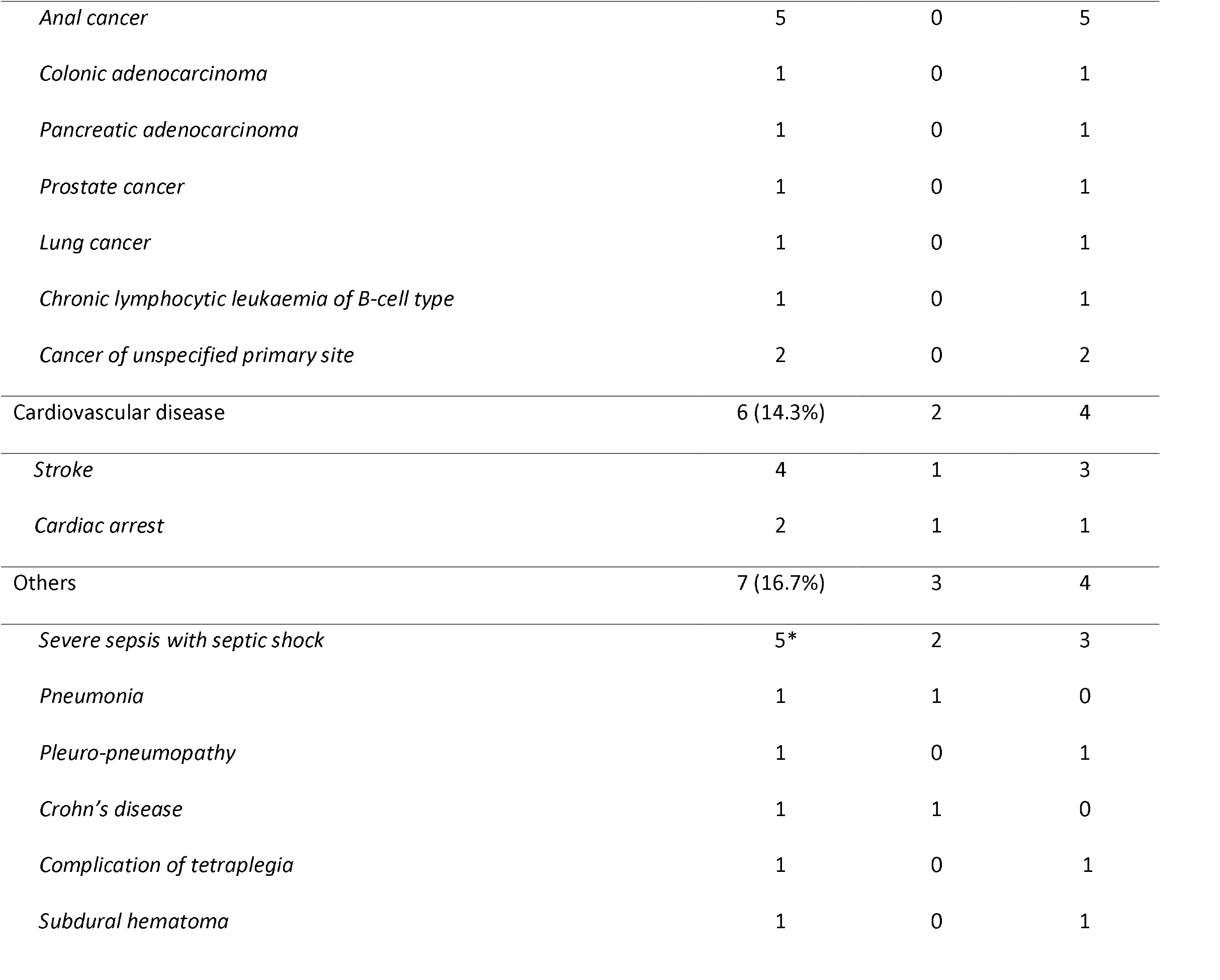
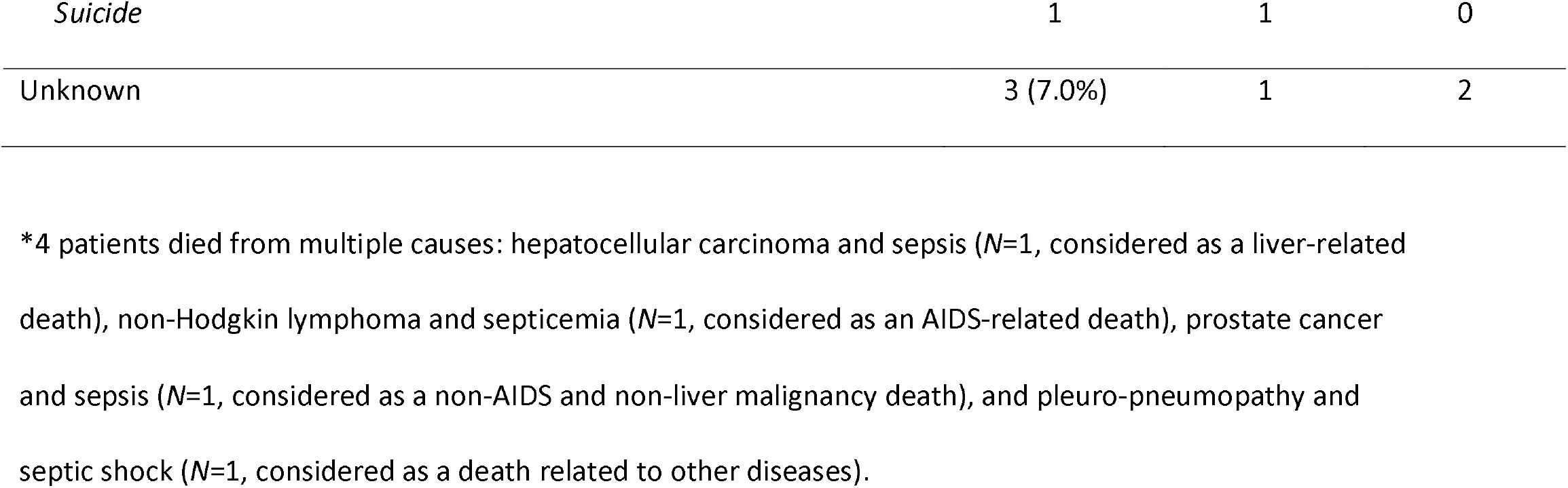
Causes of death observed in the French HIV-HBV cohort, 2002-2018.

| Cause of death | All periods<br>(N=42) | 2002-2007<br>(N=10) | 2008-2018<br>(N=32) |
| --- | --- | --- | --- |
| Liver-related disease | 7 (16.7%) | 2 | 5 |
| Hepatocellular carcinoma | 4 | 1 | 3 |
| *Intrahepatic cholangiocarcinoma* | 1 | 0 | 1 |
| *Hepatic failure* | 1 | 0 | 1 |
| *Triple infection HBV-HCV-HDV with decompensated liver disease* | 1 | 1 | 0 |
| AIDS-related disease | 7 (16.7%) | 2 | 5 |
| *Kaposi's sarcoma* | 1 | 1 | 0 |
| *Burkitt's lymphoma* | 1 | 0 | 1 |
| *Non-Hodgkin lymphoma* | 1 | 0 | 1 |
| *Progressive multifocal leukoencephalopathy* | 1 | 1 | 0 |
| *HIV-related encephalopathy* | 1 | 0 | 1 |
| *Chronic intestinal isosporiasis* | 1 | 0 | 1 |
| *HIV/AIDS disease resulting in multiple infections* | 1 | 0 | 1 |
| Non-AIDS and non-liver malignancy | 12 (28.6%) | 0 | 12 |
| *Anal cancer* | 5 | 0 | 5 |
| *Colonic adenocarcinoma* | 1 | 0 | 1 |
| *Pancreatic adenocarcinoma* | 1 | 0 | 1 |
| *Prostate cancer* | 1 | 0 | 1 |
| *Lung cancer* | 1 | 0 | 1 |
| *Chronic lymphocytic leukaemia of B-cell type* | 1 | 0 | 1 |
| *Cancer of unspecified primary site* | 2 | 0 | 2 |
| Cardiovascular disease | 6 (14.3%) | 2 | 4 |
| *Stroke* | 4 | 1 | 3 |
| *Cardiac arrest* | 2 | 1 | 1 |
| Others | 7 (16.7%) | 3 | 4 |
| *Severe sepsis with septic shock* | 5* | 2 | 3 |
| *Pneumonia* | 1 | 1 | 0 |
| *Pleuro-pneumopathy* | 1 | 0 | 1 |
| *Crohn's disease* | 1 | 1 | 0 |
| *Complication of tetraplegia* | 1 | 0 | 1 |
| *Subdural hematoma* | 1 | 0 | 1 |
| *Suicide* | 1 | 1 | 0 |
| Unknown | 3 (7.0%) | 1 | 2 |

\*4 patients died from multiple causes: hepatocellular carcinoma and sepsis ( $N=1$ , considered as a liver-related death), non-Hodgkin lymphoma and septicemia ( $N=1$ , considered as an AIDS-related death), prostate cancer and sepsis ( $N=1$ , considered as a non-AIDS and non-liver malignancy death), and pleuro-pneumopathy and septic shock ( $N=1$ , considered as a death related to other diseases).

As shown in table 2, at study entry patients who died, compared to those alive, were older (*p*<0.001), more likely to have been born in zones of low or moderate HBV prevalence (*p*<0.004), have acquired HIV infection by injecting drug use (IDU) (*p*<0.02), had other liver diseases or hepatic decompensation (*p*=0.001), had been diagnosed with an AIDS-defining event (*p*<0.001), had a longer duration of known HIV infection (*p*=0.01), lower nadir CD4+ cell counts (*p*=0.03), longer duration of ART (*p*=0.05), higher levels of AST (*p*=0.01), HBeAg-positive status (*p*=0.02), and F3-F4 fibrosis (*p*<0.001).

**Table 2.**
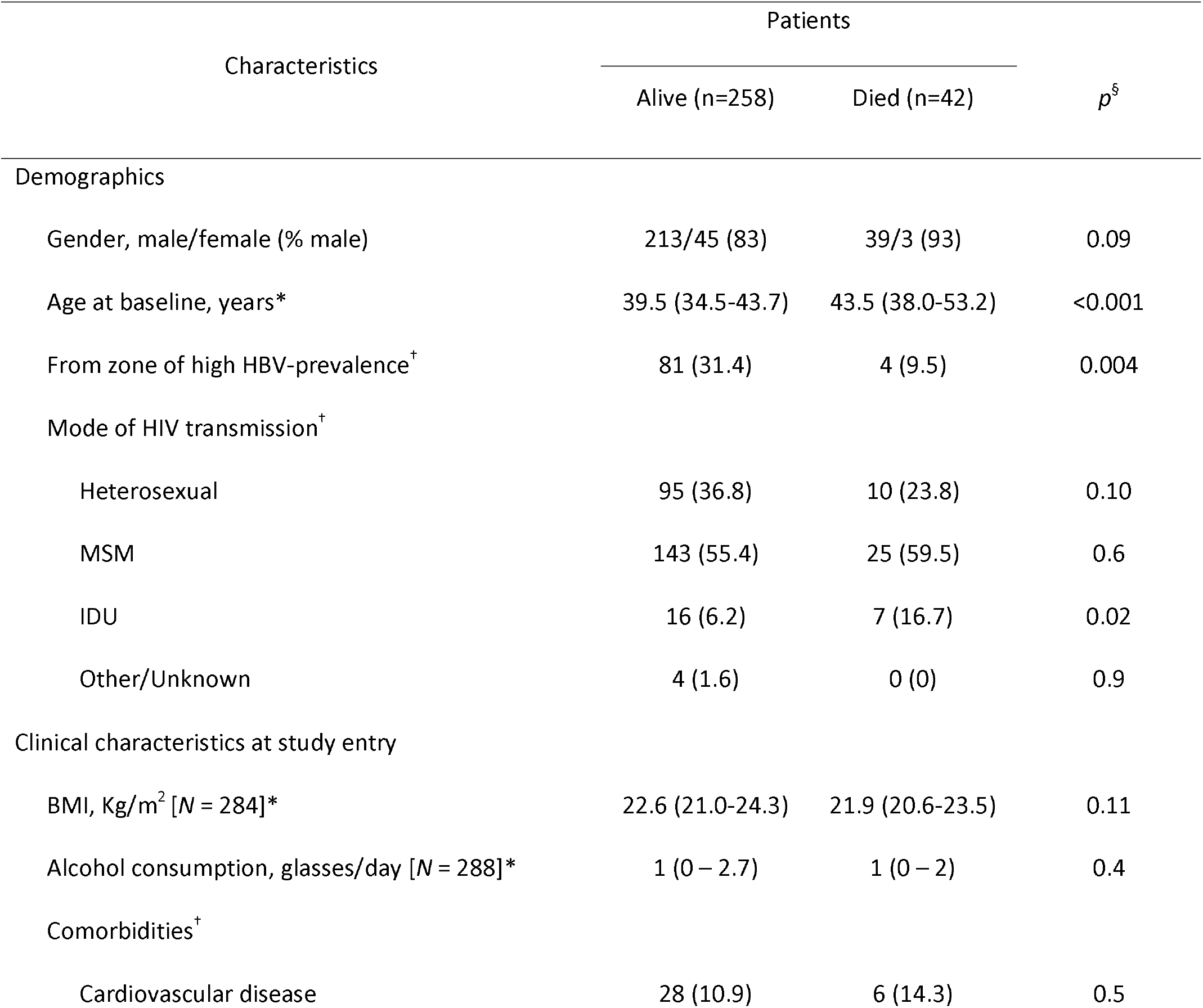
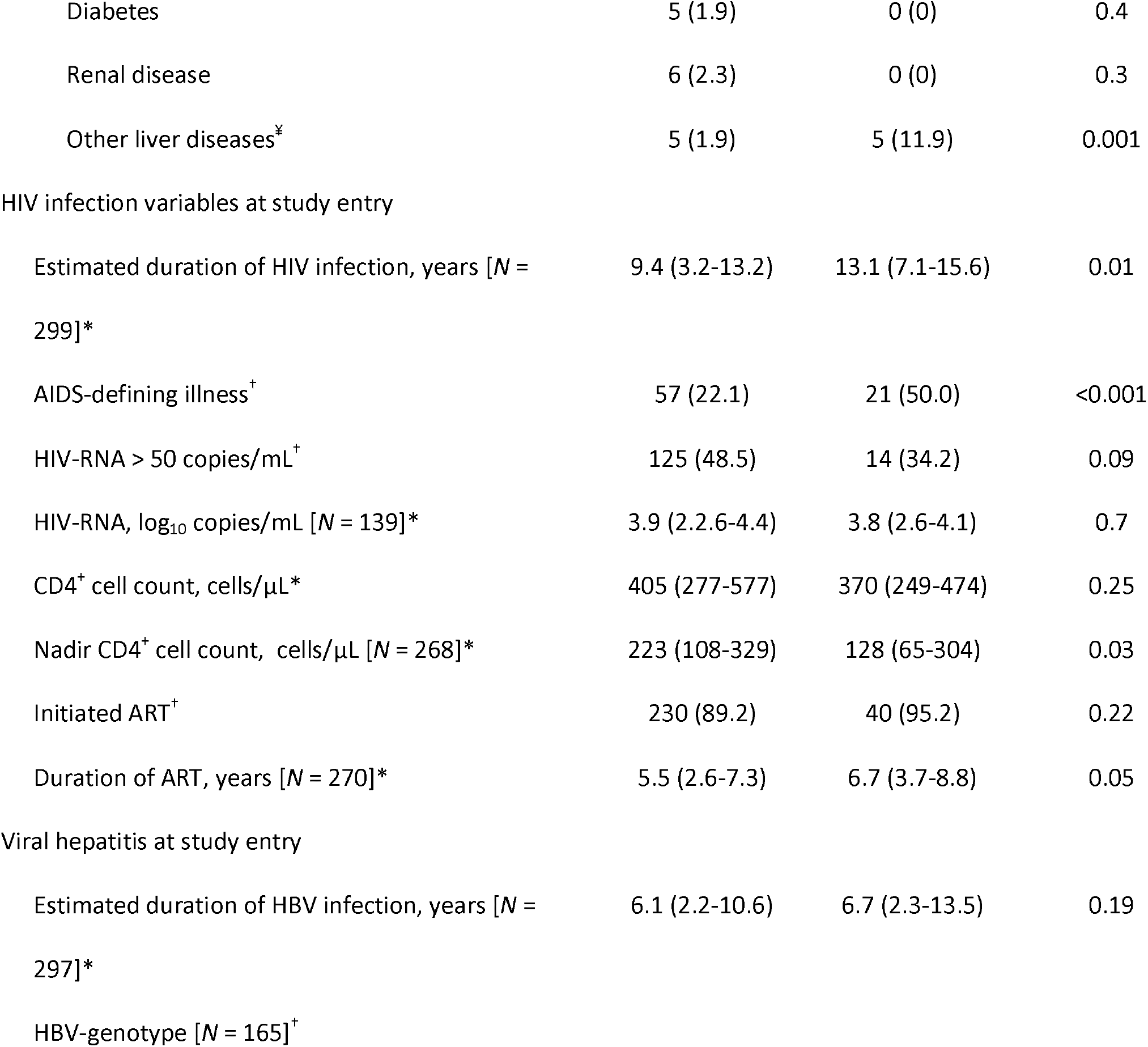
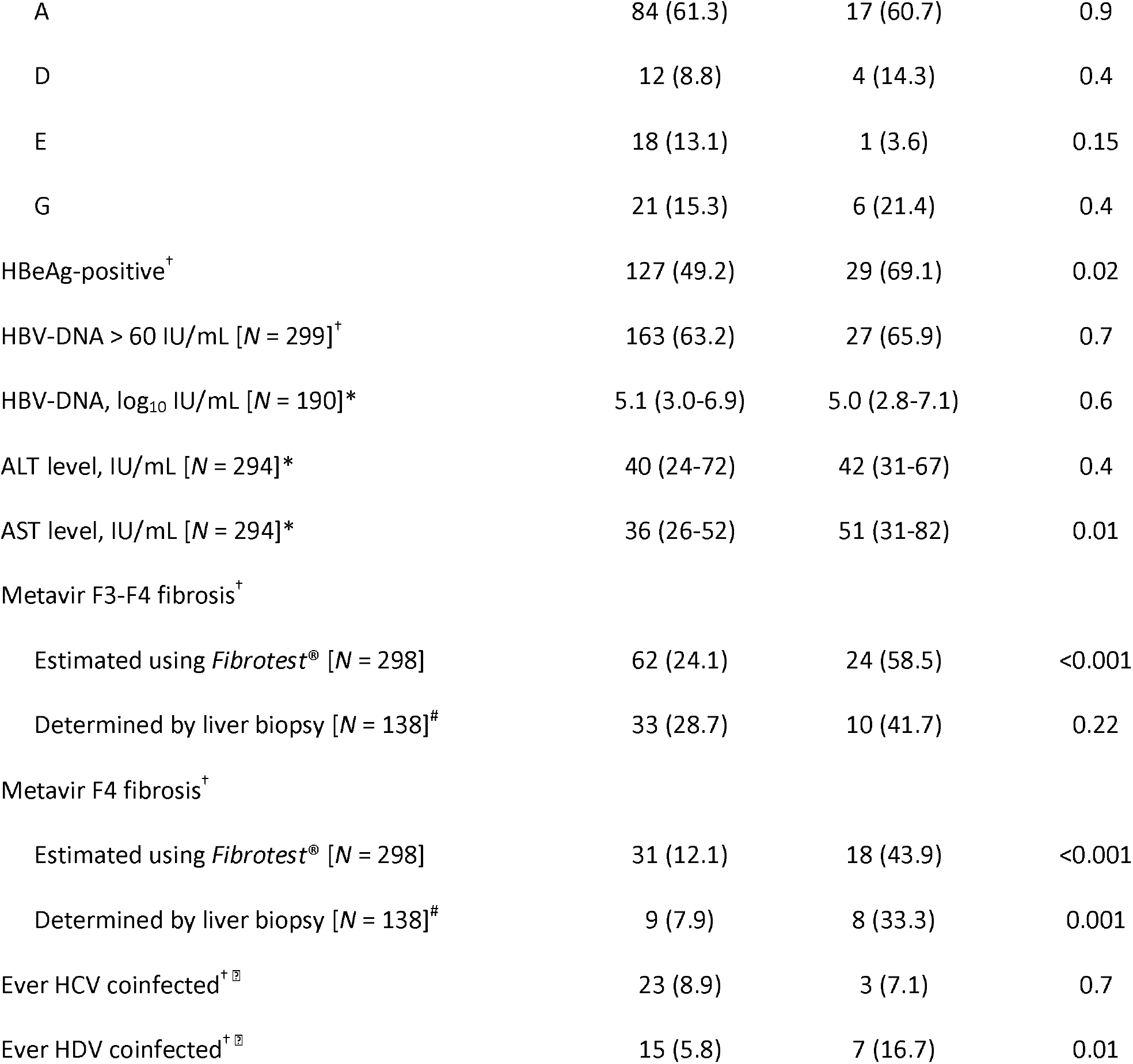
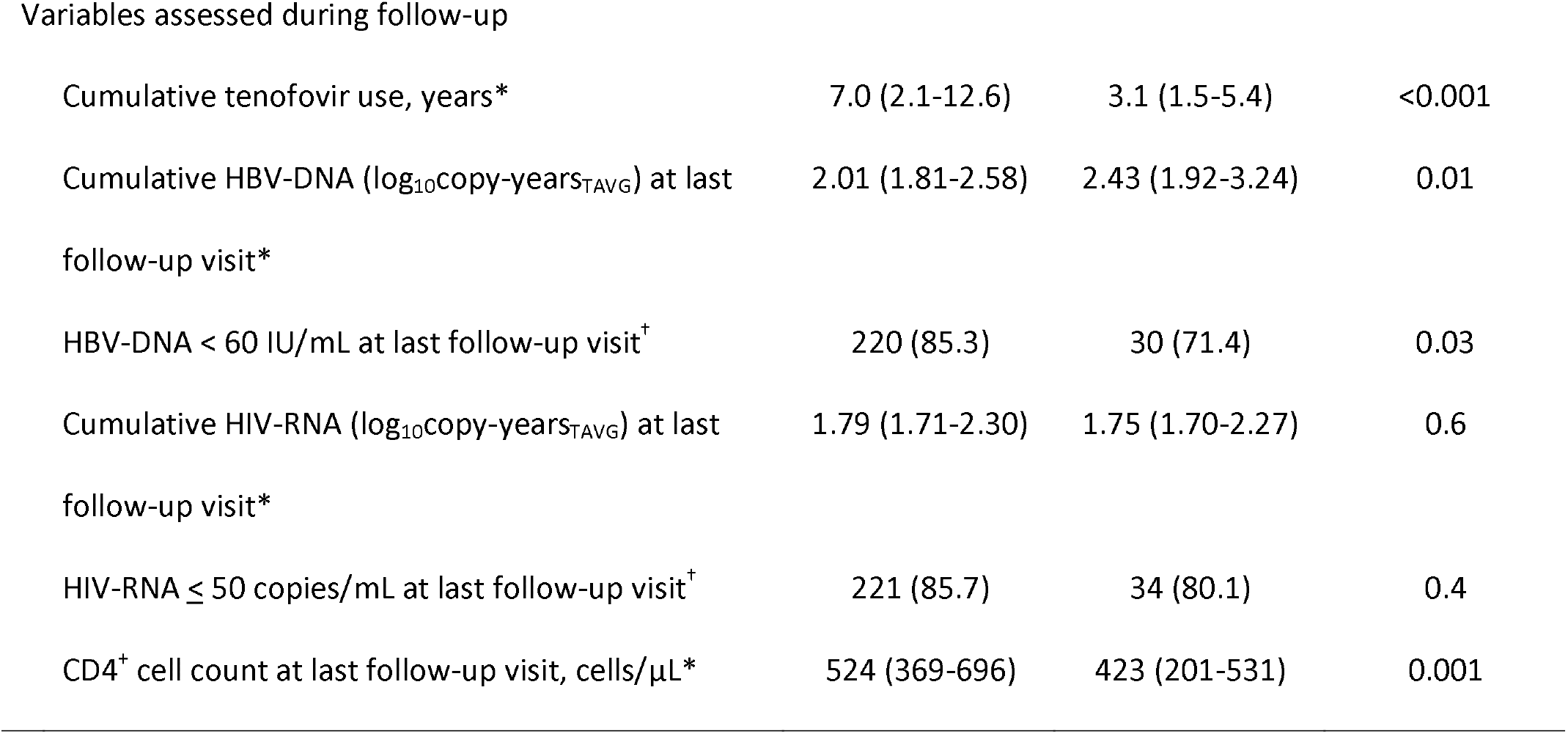
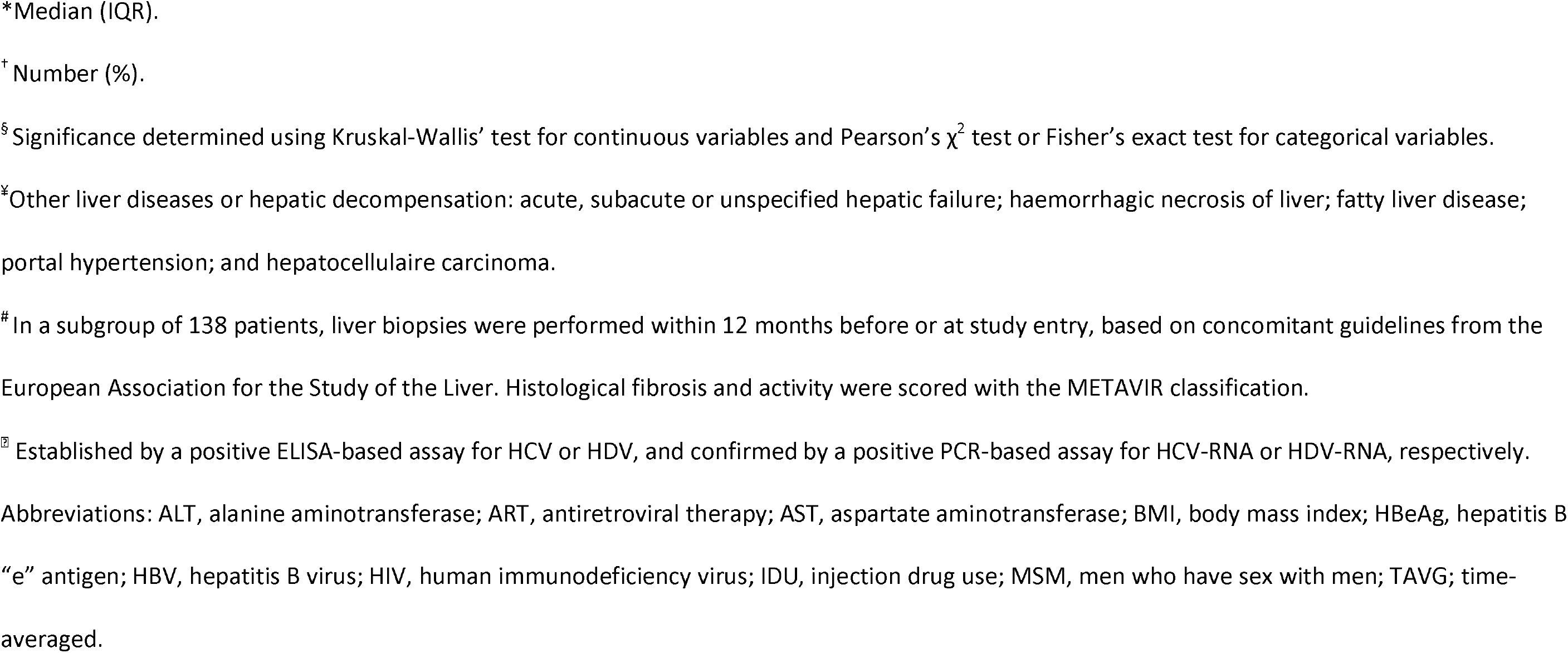
Characteristics of study population (at cohort inclusion or during follow-up)

During follow-up, patients who died, when compared to those alive, were more often diagnosed with HDV coinfection (*p*=0.01), had shorter duration of cumulative tenofovir use (*p*<0.001), lower CD4+ cell counts (*p*=0.001), and detectable HBV-DNA at last follow-up visit (p=0.03) (Table 2).

### HBV and HIV viral replication and all-cause mortality

HBV-DNA VL was more frequently detected in individuals who remained alive versus those who were deceased (Figure 2A): at 5 years, 10 years and 15 years, respectively, of 22.3% (*n*=29/130), 5.2% (*n*=7/135), 5.3% (*n*=7/133) in alive patients and 18.2% (*n*=4/22), 33.3% (*n*=2/6), 0.0% (*n*=0/1) in deceased ones (overall *p*-value<0.001). However, deceased individuals were exposed to a higher HBV-DNA level (log_10_IU/mL) at 5 years, 10 years and 15 years, respectively, of 1.29 (IQR: 1.08-1.53), 1.31 (IQR: 1.00-3.22) and 1.00 (*N*=1), compared to 1.20 (IQR: 1.08-1.78), 1.30 (IQR: 1.00-1.30) and 1.11 (IQR: 1.00-1.30) in alive patients (overall p-value<0.001) (Figure 2B). Median time-averaged copy-years of HBV-DNA (log_10_copy-years_TAVG_) between groups at 5 years, 10 years and 15 years were 2.18 (IQR: 1.85-3.10), 1.92 (IQR: 1.81-2.46), 1.88 (IQR: 1.80-2.23) for alive patients and 2.41 (IQR: 2.00-4.55), 2.49 (IQR: 2.11-3.11), 2.76 (*N*=1) for deceased ones (overall *p*-value <0.001). Conversely, both alive and deceased patients had mostly undetectable HIV-RNA VL over time (Figures 2C): at 5 years, 10 years and 15 years, respectively, 83.8% (*n*=109/130), 91.9% (*n*=124/135), 98.5% (*n*=131/133) of alive patients and 72.7% (*n*=16/22), 83.3% (*n*=5/6), 100.0% (*n*=1/1) of deceased ones (overall *p*-value=0.23). Median viral loads and time-averaged copy-years of HIV-RNA were also low throughout follow-up (Figure 2D).

**Figure 2.**
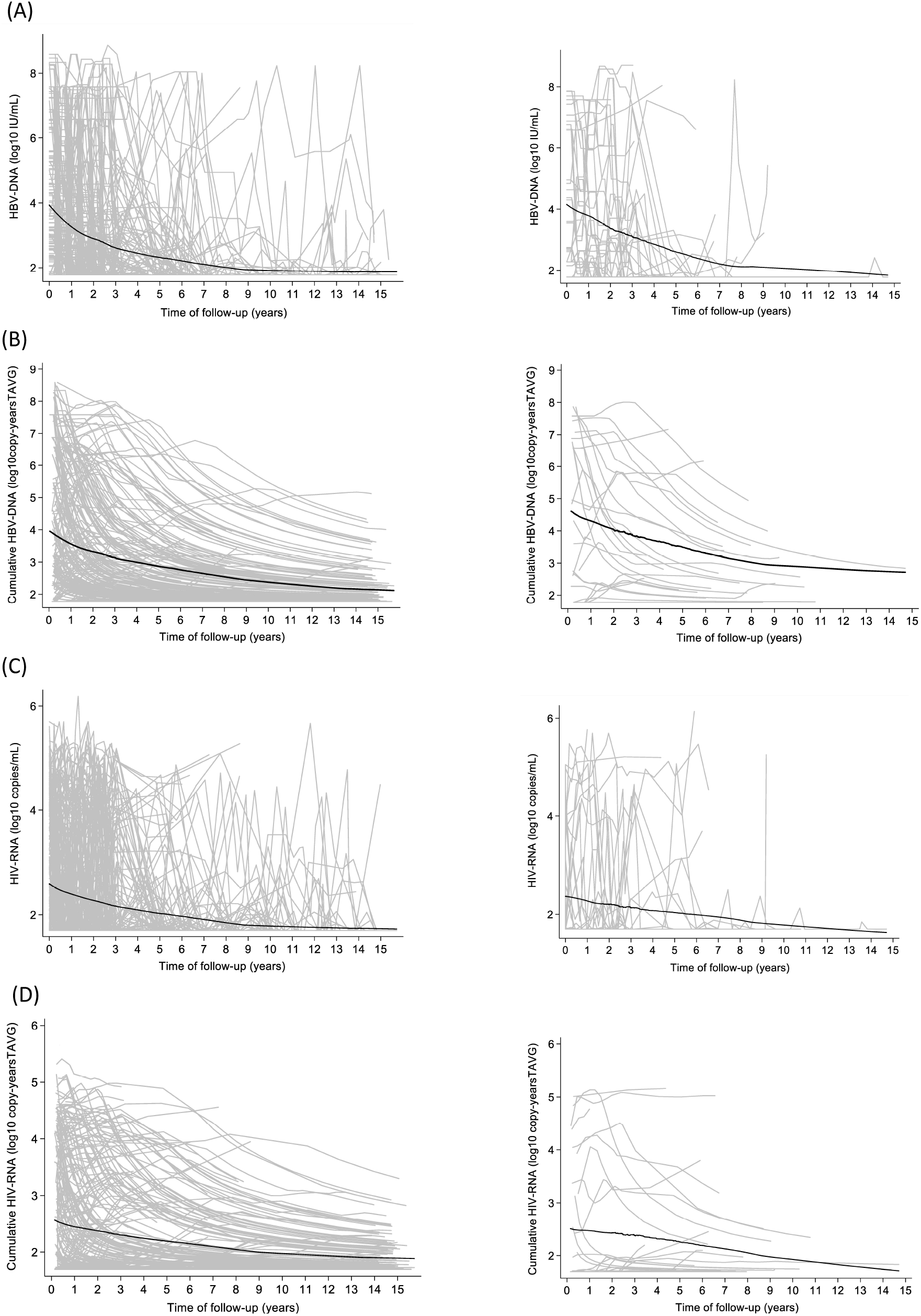
Evolution of extent and cumulative extent of viral replication over time. Evolution of (A) Hepatitis B virus (HBV)-DNA (log_10_ IU/mL), (B) cumulative HBV-DNA (log_10_ copy-years_TAVG_), (C) HIV-RNA (log_10_ copies/mL) and (D) cumulative HIV-RNA (log_10_ copy-years_TAVG_) over time according to mortality outcome (alive patients – on the left, and deceased patients – on the right). Means are expressed as bold lines from a LOWESS curve and individual levels are expressed as gray lines.

When jointly and longitudinally modeling HBV and HIV replication on all-cause mortality, we found no significant associations with undetectable HBV-DNA on mortality rates (*p*=0.08, Table 3). However, we did observe a higher rate of all-cause mortality with higher expected mean log-transformed HBV-DNA VL (*p*=0.03) and cumulative time-averaged copy-years of HBV-DNA (*p*=0.03) after adjustment for age, AIDS-defining illness and F3-F4 fibrosis level at study entry (Table 3). No significant association was found between any of the HIV-RNA outcomes and all-cause mortality (Table 3).

**Table 3.**
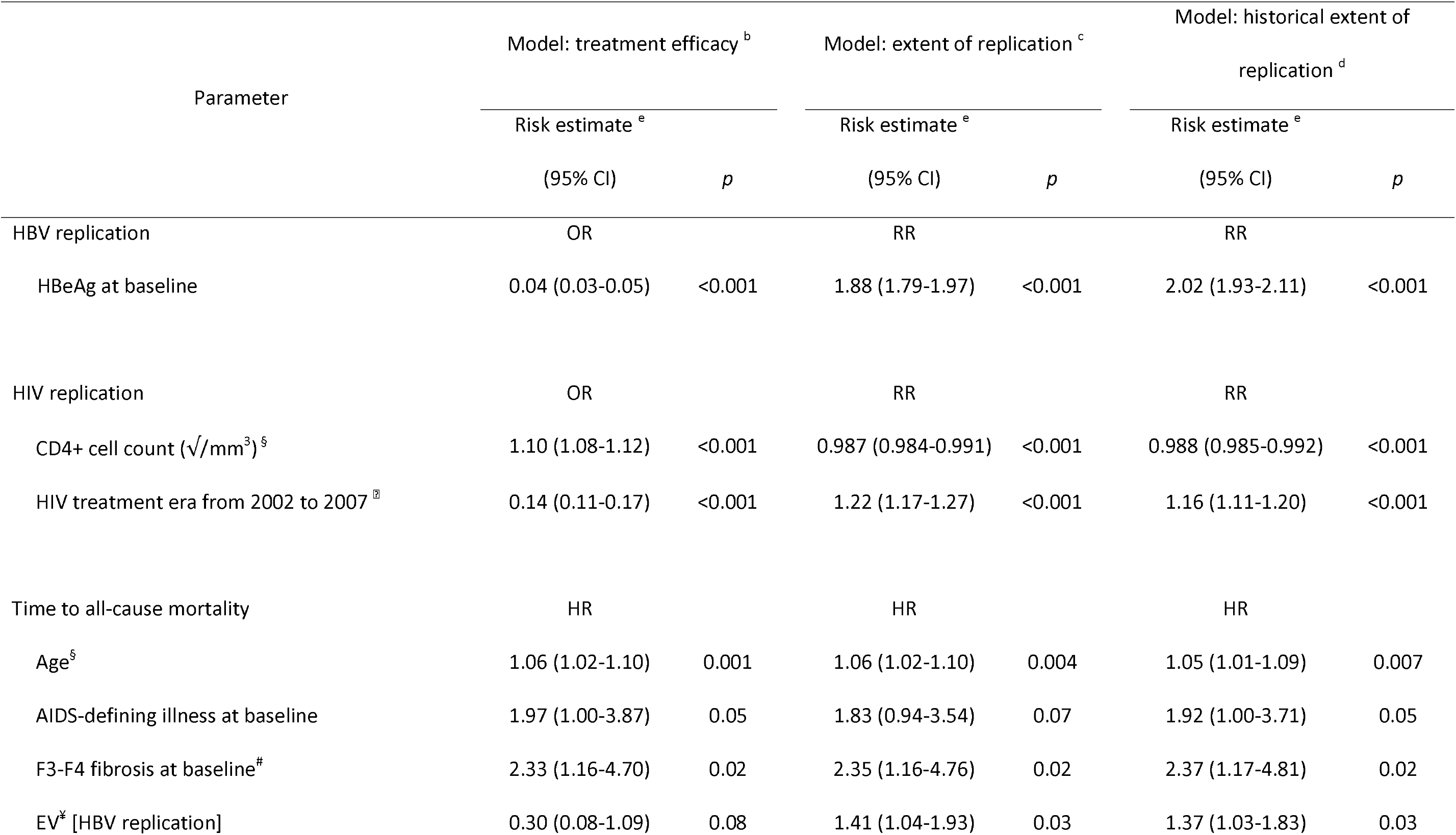
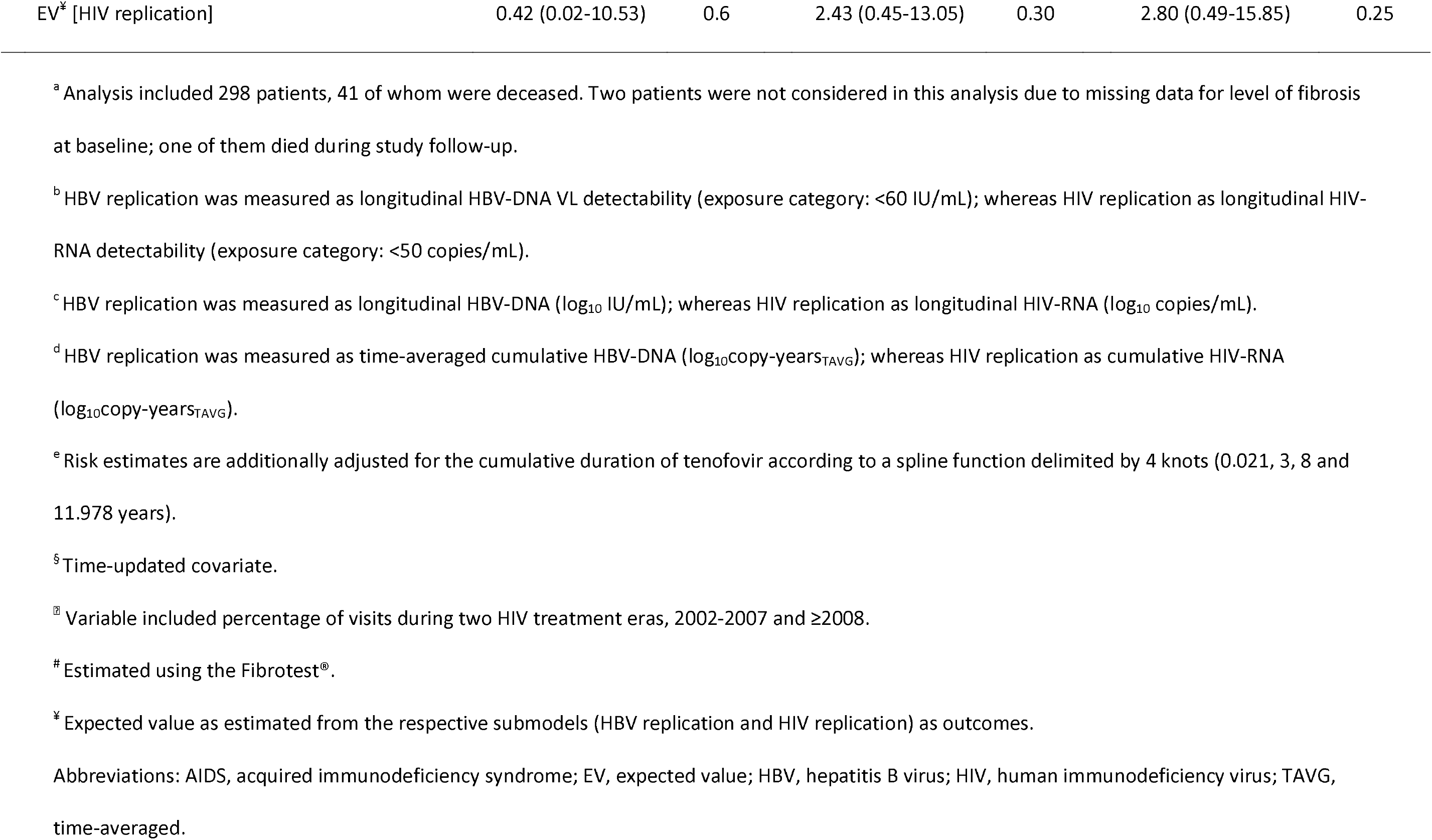
Association of HBV-DNA and HIV-RNA replication with all-cause mortality (joint models)^a^.

## Discussion

In this long-term, prospective study of treated HIV-HBV co-infected patients, the most common cause of death observed in our study was non-AIDS and non-liver-related malignancies, representing approximately one-third of deaths. Coupled with the high proportion of deaths due to CVD, the spectrum of mortality causes in HIV-HBV co-infection would appear to mirror that of aging HIV-positive individuals in general [22,23]. Nevertheless, liver-related and AIDS-related causes together represented one-third of the deaths, which occurred even during periods when use of TDF-containing ART and suppression of HIV-RNA and HBV-DNA were common at the population level.

Interestingly, the concurrent and historical extent of HBV viremia over time, but not HIV, seemed to play a major role in all-cause mortality. This effect was not observed when modeling undetectable HBV-DNA over time. Other studies in HIV-HBV co-infected patients have shown an association between higher HBV-DNA at treatment initiation and all-cause mortality [16,24]. The curious part of this finding was that this association was observed in the presence of almost exclusively TDF-treated individuals, with assumedly extensive HBV DNA suppression. Given the findings in our study, perhaps the historical exposure of high HBV DNA prior to ART initiation explained the increased risk in mortality in these studies despite effective anti-HIV and anti-HBV treatment.

Conversely, HIV-RNA had no effect on all-cause mortality in our study. Given the much more rapid on-treatment decline to HIV-RNA suppression, compared to HBV-DNA, and the improvements witnessed in overall virological response to ART with reduced pill burden, fewer side-effects, higher genetic barriers to resistance and increasing treatment options [5,25], these results could be explained by the higher degree of HIV-RNA than HBV-DNA suppression in our cohort. The carry-over effect of continuous viral suppression can be observed in the time-averaged cumulative HIV-RNA and HBV-DNA levels, where cumulative HIV-RNA levels were much lower early on during follow-up in the cohort compared to cumulative HBV-DNA. Furthermore, AIDS-related mortality is generally associated with the time spent at lower CD4+ cell counts [26]. Individuals in our cohort were on average able to attain levels of CD4+ that are associated with reduced risk of both AIDS-related and overall mortality. The low overall proportion of AIDS-related deaths is also in accordance with other findings from European cohorts of HIV-positive individuals [27].

The question is then what aspects of HBV-DNA replication are driving overall mortality. Naturally, liver-related disease due to immunological responses against prolonged HBV infection are one possibility [28]. Advanced liver fibrosis at cohort inclusion was found to be independently associated with all-cause mortality. A previous study in this cohort demonstrated nominal changes in fibrosis levels during TDF-based ART,[29] suggesting continued risk of death due to this comorbidity. Our assessment of liver fibrosis was based on a non-invasive measure with a certain degree of error;[19] however, the FibroTest® in particular has been evaluated as a tool to predict liver fibrosis evolution [30] and overall survival [31] in individuals with chronic HBV infection. Although liver fibrosis is mostly known to affect liver-related mortality (i.e., death due to HCC or end-stage liver disease), we observed few liver-related deaths. It should be noted, however, that individuals with liver cirrhosis do have a high risk of death due to invasive bacterial pathogens [32] and many of the other causes of death in our cohort were related to bacteremia. The role of bacterial pathogens in cirrhotic HIV-HBV co-infected individuals should be elucidated in larger cohorts.

One intriguing finding was that certain extra-hepatic malignant tumors caused a number of deaths in this study population. In fact, 40% of the 20 individuals who died from cancer developed an extra-hepatic tumor (i.e. anal cancer, cholangiocarcinoma, pancreatic adenocarcinoma, and non-Hodgkin lymphoma [NHL]). Previous research has found that HBsAg-positive individuals with high-risk human papillomavirus (hrHPV) had an increased risk of high-grade anal squamous intraepithelial lesions compared to those who were HBsAg-negative [33]. Furthermore, HBsAg-positive individuals exhibiting high levels of HBV activity (i.e. HBeAg-positivity and detectable HBV-DNA) have been shown to have a significantly higher risk of pancreatic carcinoma compared to HBsAg-negative individuals [34]. Finally, although NHL is normally related to immunocompromised individuals and is classified as an AIDS-defining illness, growing evidence suggests that ART-treated patients with chronic HBV infection are at increased risk for NHL [35]. How HBV activity participates in the tumorigenesis of extra-hepatic tumors is not clearly understood.

Similar to others [36], HDV-co-infected individuals had a higher all-cause mortality rate. Unfortunately, due to its close association with fibrosis, we decided not to include HDV-co-infection in multivariable analysis. Of the 22 individuals in our cohort with HIV-HBV-HDV infection, 7 died. Despite the fact that 6 of these deceased individuals had advanced liver fibrosis and at least one liver-related complication, only one died from liver-related diseases. In contrast to others [36], only one HCV-positive individual (without HDV) died in our cohort, which could be a reflection of the low proportion of injecting drug users and the increasingly effective direct acting antivirals available by the end of follow-up. Given the very few individuals with tri-/quad-infection who remained in follow-up, generalizability of our data to this population would be limited.

Our study has certain limitations. First, the study population involves HIV-positive individuals with extensive ART-experience and larger degrees of immunosuppression compared to contemporary patient populations, but still actively seen in outpatient settings. Second, HBsAg-seroclearance has been shown to reduce all-cause mortality in HBV mono-infected individuals [37], yet we had an insufficient number of events to validate this in our cohort. Third, there was a rather high rate of LTFU. Individuals who were LTFU could have had a higher risk of more advanced disease, causing non-differential LTFU bias when evaluating certain risk-factors. Population characteristics were, however, rather comparable between those who did versus did not continue follow-up [17]. Finally, we had limited to no data on alcohol use, smoking, and metabolic diseases, which could not be considered in analysis.

In conclusion, our findings provide strong evidence that HIV-HBV co-infected patients who have been exposed to HBV-DNA over time are at an elevated risk for all-cause mortality. The lack of association with HIV-RNA replication could be due to the more rapid and extensive viral suppression compared to HBV-DNA. Accompanied by the strong association between advanced liver fibrosis and overall mortality, monitoring liver fibrosis and HBV-DNA viral loads should be an essential component to help assess the prognosis of co-infected individuals. The noticeably common deaths due to extra-hepatic malignancies needs to be further studied and perhaps increased screening is called for in the HIV-HBV co-infected patient population.

## Data Availability

The datasets generated during and/or analysed during the current study are available from the corresponding author on reasonable request.

## Acknowledgements

The authors are grateful to the patients and the clinical teams for their commitment to the French HIV-HBV Cohort. This study was sponsored by the Institut de Médecine et d’Epidémiologie Appliquée (IMEA). L.N.C.D. was awarded a post-doctoral fellowship from the ANRS.

## Role of each author

L.N.C.D. was responsible for the statistical analysis, interpretation of the data, and drafting the manuscript. R.K. obtained and verified vital status on participants, assisted in the statistical analysis, and gave critical revisions of the manuscript. H.R., P.M., C. L-C., and J.C. acquired data for the cohort, assisted in interpreting data, and gave critical revisions of the manuscript. S.M., A.G. and C.D. were responsible for interpretation of the data and drafting the manuscript. K.L. helped design, conceptualize, and obtain funding for the French HIV-HBV cohort study, coordinated data collection, and drafted the manuscript. A.B. coordinated data analysis, gave important comments on data interpretation, drafted parts of the manuscript, and provided critical revisions of the manuscript. All authors have approved the final version of the article.

## References

1. Leumi S, Bigna JJ, Amougou MA, Ngouo A, Nyaga UF, Noubiap JJ. Global burden of hepatitis B infection in people living with human immunodeficiency virus: a systematic review and meta-analysis. Clin Infect Dis 2020; 71:2799–2806.

2. Falade-Nwulia O, Seaberg EC, Rinaldo CR, Badri S, Witt M, Thio CL. Comparative risk of liver-related mortality from chronic hepatitis B versus chronic hepatitis C virus infection. Clin Infect Dis 2012; 55:507–513.

3. Mellors JW, Muñoz A, Giorgi JV, et al. Plasma viral load and CD4+ lymphocytes as prognostic markers of HIV-1 infection. Ann Intern Med 1997; 126:946–954.

4. Antiretroviral Therapy Cohort Collaboration (ART-CC), Lanoy E, May M, et al. Prognosis of patients treated with cART from 36 months after initiation, according to current and previous CD4 cell count and plasma HIV-1 RNA measurements. AIDS Lond Engl 2009; 23:2199–2208.

5. Smith CJ, Ryom L, Weber R, et al. Trends in underlying causes of death in people with HIV from 1999 to 2011 (D:A:D): a multicohort collaboration. The Lancet 2014; 384:241–248.

6. Iloeje UH, Yang H, Jen C, et al. Risk and predictors of mortality associated with chronic hepatitis B infection. Clin Gastroenterol Hepatol 2007; 5:921–931.

7. Marcellin P, Gane E, Buti M, et al. Regression of cirrhosis during treatment with tenofovir disoproxil fumarate for chronic hepatitis B: a 5-year open-label follow-up study. Lancet Lond Engl 2013; 381:468–475.

8. Papatheodoridis GV, Chan HL-Y, Hansen BE, Janssen HLA, Lampertico P. Risk of hepatocellular carcinoma in chronic hepatitis B: assessment and modification with current antiviral therapy. J Hepatol 2015; 62:956–967.

9. Boyd A, Gozlan J, Maylin S, et al. Persistent viremia in human immunodeficiency virus/hepatitis B coinfected patients undergoing long-term tenofovir: Virological and clinical implications. Hepatology 2014; 60:497–507.

10. Huang Y-S, Cheng C-Y, Liou B-H, et al. Efficacy and safety of elvitegravir/cobicistat/emtricitabine/tenofovir alafenamide as maintenance treatment in HIV/HBV-coinfected patients. J Acquir Immune Defic Syndr 2021; 86:473–481.

11. European AIDS Clinical Society. Guidelines Version 10.1. October 2020.

12. van Welzen BJ, Smit C, Boyd A, et al. Decreased all-cause and liver-related mortality risk in HIV/hepatitis B virus coinfection coinciding with the introduction of tenofovir-containing combination antiretroviral therapy. Open Forum Infect Dis 2020; 7:ofaa226.

13. Tsai W-C, Hsu W-T, Liu W-D, et al. Impact of antiretroviral therapy containing tenofovir disoproxil fumarate on the survival of patients with HBV and HIV coinfection. Liver Int Off J Int Assoc Study Liver 2019; 39:1408–1417.

14. Wandeler G, Mauron E, Atkinson A, et al. Incidence of hepatocellular carcinoma in HIV/HBV-coinfected patients on tenofovir therapy: Relevance for screening strategies. J Hepatol 2019; 71:274–280.

15. Christian B, Fabian E, Macha I, et al. Hepatitis B virus coinfection is associated with high early mortality in HIV-infected Tanzanians on antiretroviral therapy. AIDS Lond Engl 2019; 33:465–473.

16. Kouamé G-M, Boyd A, Moh R, et al. Higher mortality despite early antiretroviral therapy in human immunodeficiency virus and hepatitis B virus (HBV)-coinfected patients with high HBV replication. Clin Infect Dis Off Publ Infect Dis Soc Am 2018; 66:112–120.

17. Boyd A, Dezanet LNC, Kassime R, et al. Subclinical and clinical outcomes in HIV and chronic hepatitis B virus co-infected patients from clinical outpatient centers in France: an ambi-spective longitudinal cohort study. JMIR 2020 (In press). Available at: https://preprints.jmir.org/preprint/24731.

18. Poynard T, Ngo Y, Munteanu M, Thabut D, Ratziu V. Noninvasive markers of hepatic fibrosis in chronic hepatitis B. Curr Hepat Rep 2011; 10:87–97.

19. Bottero J, Lacombe K, Guéchot J, et al. Performance of 11 biomarkers for liver fibrosis assessment in HIV/HBV co-infected patients. J Hepatol 2009; 50:1074–1083.

20. Boyd A, Gozlan J, Miailhes P, et al. Rates and determinants of hepatitis B ‘e’ antigen and hepatitis B surface antigen seroclearance during long-term follow-up of patients coinfected with HIV and hepatitis B virus. AIDS 2015; 29:1963–1973.

21. Crowther MJ. merlin—A unified modeling framework for data analysis and methods development in Stata. Stata J Promot Commun Stat Stata 2020; 20:763–784.

22. Joint United Nations Programme on HIV/AIDS. HIV and aging. 2013. Available at: http://www.unaids.org/en/media/unaids/contentassets/documents/unaidspublication/2013/20131101_JC2563_hiv-and-aging_en.pdf. Accessed 19 November 2020.

23. Mary-Krause M, Grabar S, Lievre L, et al. Cohort profile: French hospital database on HIV (FHDH-ANRS CO4). Int J Epidemiol 2014; 43:1425–1436.

24. Velen K, Charalambous S, Innes C, Churchyard GJ, Hoffmann CJ. Chronic hepatitis B increases mortality and complexity among HIV-coinfected patients in South Africa: a cohort study. HIV Med 2016; 17:702–707.

25. Trickey A, May MT, Vehreschild J-J, et al. Survival of HIV-positive patients starting antiretroviral therapy between 1996 and 2013: a collaborative analysis of cohort studies. Lancet HIV 2017; 4:e349–e356.

26. Battegay M, Nüesch R, Hirschel B, Kaufmann GR. Immunological recovery and antiretroviral therapy in HIV-1 infection. Lancet Infect Dis 2006; 6:280–287.

27. Vandenhende M-A, Roussillon C, Henard S, et al. Cancer-related causes of death among HIV-infected patients in France in 2010: evolution since 2000. PloS One 2015; 10:e0129550.

28. Singh KP, Crane M, Audsley J, Avihingsanon A, Sasadeusz J, Lewin SR. HIV-hepatitis B virus coinfection: epidemiology, pathogenesis, and treatment. AIDS 2017; 31:2035–2052.

29. Boyd A, Bottero J, Miailhes P, et al. Liver fibrosis regression and progression during controlled hepatitis B virus infection among HIV-HBV patients treated with tenofovir disoproxil fumarate in France: a prospective cohort study. J Int AIDS Soc 2017; 20:21426.

30. Poynard T, Munteanu M, Deckmyn O, et al. Validation of liver fibrosis biomarker (FibroTest) for assessing liver fibrosis progression: proof of concept and first application in a large population. J Hepatol 2012; 57:541–548.

31. de Lédinghen V, Vergniol J, Barthe C, et al. Non-invasive tests for fibrosis and liver stiffness predict 5-year survival of patients chronically infected with hepatitis B virus. Aliment Pharmacol Ther 2013; 37:979–988.

32. Nahon P, Lescat M, Layese R, et al. Bacterial infection in compensated viral cirrhosis impairs 5-year survival (ANRS CO12 CirVir prospective cohort). Gut 2017; 66:330–341.

33. McCloskey JC, Kast WM, Flexman JP, McCallum D, French MA, Phillips M. Syndemic synergy of HPV and other sexually transmitted pathogens in the development of high-grade anal squamous intraepithelial lesions. Papillomavirus Res Amst Neth 2017; 4:90–98.

34. Iloeje UH, Yang H-I, Jen C-L, et al. Risk of pancreatic cancer in chronic hepatitis B virus infection: data from the REVEAL-HBV cohort study. Liver Int 2010; 30:423–429.

35. Wang Q, De Luca A, Smith C, et al. Chronic hepatitis B and C virus infection and risk for non-Hodgkin lymphoma in HIV-infected patients: a cohort study. Ann Intern Med 2017; 166:9.

36. Béguelin C, Moradpour D, Sahli R, et al. Hepatitis delta-associated mortality in HIV/HBV-coinfected patients. J Hepatol 2017; 66:297–303.

37. Arase Y, Ikeda K, Suzuki F, et al. Long-term outcome after hepatitis B surface antigen seroclearance in patients with chronic hepatitis B. Am J Med 2006; 119:71.e9–16.

