## Supplementary Table 1 for "Effect of viral replication and liver fibrosis on all-cause mortality in HIV/HBV coinfected patients: a retrospective analysis of a 15-year longitudinal cohort"

Supplementary Table 1 - Risk factors for all-cause mortality.

| Characteristics | | | Univariate | |  | Multivariate^a^ | |
| --- | --- | --- | --- | --- | --- | --- | --- |
|  |  |  | HR (CI 95%) | *p* |  | HR (CI 95%) | *p* |
| Demographics | | |  |  |  |  |  |
|  | Gender, male/female (% male) | | 2.62 (0.81-8.47) | 0.11 |  |  |  |
|  | Age at baseline, years | | 1.07 (1.04-1.11) | <0.001 |  | 1.05 (1.01-1.09) | 0.01 |
|  | From zone of high HBV-prevalence | | 0.25 (0.09-0.71) | 0.009 |  |  |  |
| Clinical characteristics at study entry | | |  |  |  |  |  |
|  | BMI, Kg/m^2^ | | 0.92 (0.82-1.03) | 0.15 |  |  |  |
|  | Alcohol consumption, glasses/day | | 1.03 (0.89-1.21) | 0.7 |  |  |  |
| HIV infection variables at study entry | | |  |  |  |  |  |
|  | Estimated duration of HIV infection, years | | 1.08 (1.02-1.14) | 0.01 |  |  |  |
|  | AIDS-defining illness | | 2.99 (1.63-5.47) | <0.001 |  | 2.35 (1.25-4.41) | 0.008 |
|  | CD4^+^ cell count, per 100 cells/µL | | 0.92 (0.81-1.05) | 0.23 |  |  |  |
|  | Nadir CD4^+^ cell count, per 100 cells/µL | | 0.80 (0.64-1.00) | 0.05 |  |  |  |
|  | Initiated ART | | 1.08 (0.99-1.17) | 0.08 |  |  |  |
|  | Duration of ART, years | | 1.03 (0.98-1.09) | 0.25 |  |  |  |
| Viral hepatitis | | |  |  |  |  |  |
|  | Estimated duration of HBV infection, years | | 1.03 (0.98-1.09) | 0.22 |  |  |  |
|  | HBeAg-positive | | 2.07 (1.08-3.99) | 0.03 |  |  |  |
|  | ALT level, IU/mL | | 1.00 (0.99-1.00) | 0.8 |  |  |  |
|  | AST level, IU/mL | | 1.008 (1.002-1.01) | 0.01 |  |  |  |
|  | Metavir F3-F4 fibrosis^†^ | |  |  |  |  |  |
|  |  | Estimated using *Fibrotest*® | 3.48 (1.87-6.48) | <0.001 |  | 2.12 (1.06-4.21) | 0.03 |
|  |  | Determined by liver biopsy ^#^ | 1.70 (1.18-2.45) | 0.005 |  |  |  |
|  | Ever HCV coinfected | | 0.28 (0.04-2.01) | 0.20 |  |  |  |
|  | Ever HDV coinfected | | 3.13 (1.39-7.04) | 0.006 |  |  |  |
| Variables assessed during the follow-up study | | |  |  |  |  |  |
|  | Cumulative tenofovir use, years | | 0.98 (0.91-1.06) | 0.6 |  |  |  |
|  | Time-updated CD4^+^ cell count, cells/µL | | 0.997 (0.996-0.999) | 0.003 |  |  |  |
|  | CD4^+^ cell count at last follow-up visit, cells/µL | | 0.996 (0.995-0.998) | <0.001 |  |  |  |
|  | Time-updated F3-F4 fibrosis level | | 4.48 (1.43-14.08) | 0.01 |  |  |  |

^a^ In multivariate modeling, BMI had too many missing data and was not considered further; AST levels, HDV coinfection, F3-F4 fibrosis (estimated using FibroTest) and time-updated F3-F4 fibrosis levels were collinear and we preferred F3-F4 fibrosis (estimated using FibroTest); CD4^+^ cell count at the last follow-up visit, initiation of ART and nadir CD4^+^ cell count were collinear and we preferred nadir CD4^+^ cell count; HBeAg status and time-updated CD4^+^ cell count were included in the submodels of the joint models analysis and were not considered further. The following variables were removed as their association was no longer significant in the multivariable model: male gender (p=0.6), from zone of high prevalence (p=0.17), and nadir CD4^+^ cell count (p=0.4). The final multivariate model was adjusted by all covariates listed.

^#^ In a subgroup of 138 patients, liver biopsies were performed within 12 months before or at study entry, based on concomitant guidelines from the European Association for the Study of the Liver. Histological fibrosis and activity were scored with the METAVIR classification.

Abbreviations: ALT, alanine aminotransferase; ART, antiretroviral therapy; AST, aspartate aminotransferase; BMI, body mass index; HBeAg, hepatitis B “e” antigen; HBV, hepatitis B virus; HIV, human immunodeficiency virus; IDU, injection drug use; MSM, men who have sex with men; ULN, upper limit of normality.
